# Germline TYK2 mutation and cancer risk

**DOI:** 10.1101/2022.01.26.22269888

**Authors:** James Yarmolinsky, Christopher I. Amos, Rayjean J. Hung, Victor Moreno, Kimberley Burrows, Karl Smith-Byrne, Joshua R. Atkins, Paul Brennan, Colon Cancer Family Registry (CCFR), Colorectal Cancer Transdisciplinary study (CORECT), Genetics and Epidemiology of Colorectal Cancer Consortium (GECCO), Prostate Cancer Association Group to Investigate Cancer Associated Alterations in the Genome (PRACTICAL) consortium, James D. McKay, Richard M. Martin, George Davey Smith

**Author notes:** **Corresponding author:** James Yarmolinsky, PhD, MRC Integrative Epidemiology Unit, Population Health Sciences, Bristol Medical School, University of Bristol, Bristol, UK. Members of the Colon Cancer Family Registry (CCFR), Colorectal Cancer Transdisciplinary study (CORECT), Genetics and Epidemiology of Colorectal Cancer Consortium (GECCO), and Prostate Cancer Association Group to Investigate Cancer Associated Alterations in the Genome (PRACTICAL) consortium are provided in **Supplementary Material**. Further information on the PRACTICAL consortium can be found at http://practical.icr.ac.uk/.

## Abstract

Deucravacitinib, a novel, selective inhibitor of TYK2 is currently under review at the FDA and EMA for treatment of moderate-to-severe plaque psoriasis. It is unclear whether recent safety concerns (i.e. elevated rates of lung cancer and lymphoma) related to similar medications (i.e. other JAK inhibitors) are shared with this novel TYK2 inhibitor. We used a partial loss-of-function variant in TYK2 (rs34536443), previously shown to protect against psoriasis and other autoimmune diseases, to evaluate the potential effect of therapeutic TYK2 inhibition on risk of lung cancer and non-Hodgkin lymphoma. Summary genetic association data on lung cancer risk were obtained from a GWAS meta-analysis of 29,266 cases and 56,450 controls in the Integrative Analysis of Lung Cancer Risk and Etiology (INTEGRAL) consortium. Summary genetic association data on non-Hodgkin lymphoma risk were obtained from a GWAS meta-analysis of 8,489 cases and 374,506 controls in the UK Biobank and InterLymph consortium. In the primary analysis, each copy of the minor allele of rs34536443, representing partial TYK2 inhibition, was associated with an increased risk of lung cancer (OR 1.15, 95% CI 1.07-1.24, *P* = 1.72 × 10^-4^) and non-Hodgkin lymphoma (OR 1.18, 95% CI 1.05-1.33, *P* = 5.25 × 10^-3^). In secondary analyses, there was weak evidence of an association of rs34536443 with advanced prostate cancer risk (OR 1.08, 95% CI 1.00-1.17, *P* = 0.04), but little evidence of association with three other common adult cancers examined. Our analyses using an established partial loss-of-function mutation to mimic TYK2 inhibition provide genetic evidence that therapeutic TYK2 inhibition may increase risk of lung cancer and non-Hodgkin lymphoma. These findings, consistent with recent reports from post-marketing trials of similar JAK inhibitors, could have important implications for future safety assessment of Deucravacitinib and other TYK2 inhibitors in development.

## Background

On September 1, 2021 the U.S. Food and Drug Administration (FDA) announced that three Janus kinase (JAK) inhibitors approved to treat chronic inflammatory conditions would require safety warnings over increased rates of serious heart-related events, lung cancer, and lymphoma^1^. Recently, oral Deucravacitinib, a selective inhibitor of Tyrosine kinase 2 (TYK2, a member of the JAK family), was shown to lead to larger improvements in symptom severity for patients with moderate-to-severe plaque psoriasis than oral Apremilast, the current standard of care^2,3^. Deucravacitinib, therefore, has the potential to become an important treatment option for patients with psoriasis requiring systemic treatment and is currently under review for approval at the FDA and European Medicines Agency^2^. It is unclear, however, whether the elevated cancer risk associated with some JAK inhibitors is shared with this novel TYK2 inhibitor.

In the absence of long-term clinical trial data, naturally occurring genetic variation can be leveraged to validate therapeutic targets and predict their adverse effects^4^. Specifically, germline mutations causing partial or complete loss-of-function (LOF) of genes encoding drug targets can be employed to mimic pharmacological inhibition of these targets and have been used to correctly predict adverse effects of new medications^5^.

Here, we used an established partial LOF mutation in *TYK2* (rs34536443), previously shown to protect against psoriasis and other autoimmune diseases^6,7^, to evaluate the potential effect of therapeutic TYK2 inhibition on risk of lung cancer and non-Hodgkin lymphoma.

## Methods

Minor allele homozygosity of rs34536443 causes near-complete (∼80%) loss of TYK2 function, while heterozygotes have a more modest reduction in function (<40%), suggesting non-additive effects of this variant^6^. To validate this variant as a surrogate for therapeutic TYK2 inhibition, we evaluated the effect of each copy of the minor allele of rs34536443, representing partial TYK2 inhibition, on risk of psoriasis, inflammatory bowel disease, Crohn’s disease, and multiple sclerosis. These analyses were performed using summary genetic association data on up to 78,334 cases and 150,030 controls from genome-wide association studies (GWAS) of these autoimmune diseases^8-11^.

To evaluate the effect of this variant on risk of overall and histological subtype-specific lung cancer we obtained summary genetic association data on up to 29,266 cases and 56,450 controls from a GWAS meta-analysis of the Integrative Analysis of Lung Cancer Risk and Etiology (INTEGRAL)^12^. Summary genetic association data for non-Hodgkin lymphoma (NHL) were generated by meta-analysing genome-wide association (GWAS) data for rs34536443 from the UK Biobank and InterLymph consortium in METAL^13^.

In secondary analyses, we explored whether rs34536443 increased risk of three other common adult cancers (breast, colorectal, prostate), which, along with lung cancer, account for approximately half of all new U.S. cancer cases^14^. Summary genetic association data on overall and histological subtype-specific breast, colorectal, and prostate cancer on up to 260,346 cases and 234,774 controls were obtained from analyses of the Breast Cancer Association Consortium (BCAC), Genetics and Epidemiology of Colorectal Cancer Consortium (GECCO), ColoRectal Cancer Transdisciplinary Study (CORECT), Colon Cancer Family Registry (CCFR), and the Prostate Cancer Association Group to Investigate Cancer Associated Alterations in the Genome (PRACTICAL) consortium^15-17^.

All analyses were restricted to participants of European ancestry. Further information on statistical analysis, imputation, and quality control measures for these studies is provided in **Supplementary Material** and in the original publications. All studies contributing data to these analyses had the relevant institutional review board approval from each country, in accordance with the Declaration of Helsinki, and all participants provided informed consent.

## Results

To validate rs34536443 as a surrogate for therapeutic TYK2 inhibition, we confirmed that each copy of the minor allele, representing partial TYK2 inhibition, was associated with lower risk of psoriasis (OR 0.48, 95% CI 0.39-0.58, *P* = 9.01 × 10^-14^) and other autoimmune diseases (**Table 1**).

**Table 1.** Effect of the minor allele of TYK2 variant rs34536443 on risk of autoimmune conditions, lung cancer and non-Hodgkin lymphoma, and other common adult cancers

| **Outcome** | **N (cases, controls)** | **OR (95% CI)** | **P-value** |
| --- | --- | --- | --- |
| *Autoimmune conditions* |  |  |  |
| Psoriasis | 10,588; 22,806 | 0.48 (0.39-0.58) | $9.01 \times 10^{-14}$ |
| Crohn's disease | 5,956; 14,927 | 0.65 (0.55-0.76) | $3.29 \times 10^{-7}$ |
| Multiple sclerosis | 47,429; 68,374 | 0.77 (0.69-0.85) | $2.87 \times 10^{-7}$ |
| Rheumatoid arthritis | 14,361; 43,923 | 0.68 (0.63-0.75) | $4.60 \times 10^{-16}$ |
| *Primary cancer outcomes* |  |  |  |
| Lung cancer | 29,266; 56,450 | 1.15 (1.07-1.24) | $1.72 \times 10^{-4}$ |
| Current or former smokers | 23,223; 16,964 | 1.15 (1.09-1.23) | $2.29 \times 10^{-6}$ |
| Never smokers | 2,355; 7,504 | 1.09 (0.91-1.32) | 0.34 |
| Lung adenocarcinoma | 11,273; 55,483 | 1.17 (1.08-1.27) | $1.37 \times 10^{-4}$ |
| Squamous cell carcinoma | 7,426; 55,627 | 1.19 (1.08-1.31) | $2.98 \times 10^{-4}$ |
| Small cell lung cancer | 2,664; 21,444 | 1.16 (1.00-1.34) | 0.05 |
| Non-Hodgkin lymphoma | 8,489; 374,506 | 1.18 (1.05-1.33) | $5.25 \times 10^{-3}$ |
| *Secondary cancer outcomes* |  |  |  |
| Breast cancer | 122,977; 105,974 | 0.99 (0.96-1.03) | 0.69 |
| ER+ Breast cancer | 69,501; 105,974 | 1.00 (0.96-1.04) | 0.95 |
| ER- Breast cancer | 21,468; 105,974 | 1.00 (0.96-1.04) | 0.96 |
| Colorectal cancer | 58,221; 67,694 | 1.03 (0.99-1.08) | 0.18 |
| Colon cancer | 32,002; 64,159 | 1.02 (0.97-1.08) | 0.39 |
| Rectal cancer | 16,212; 64,159 | 1.05 (0.98-1.13) | 0.15 |
| Prostate cancer | 79,148; 61,106 | 1.04 (1.00-1.09) | 0.07 |
| Advanced prostate cancer | 15,167; 58,308 | 1.08 (1.00-1.17) | 0.04 |

OR represents the exponential change in odds of cancer per each copy of the minor allele of rs34536443.
Advanced prostate cancer defined as Gleason score $\geq 8$ , prostate-specific antigen > 100 ng/mL, metastatic disease (M1), or death from prostate cancer.

In analyses of 29,266 lung cancer cases and 56,540 controls, each copy of the minor allele of rs34536443 was associated with an increased risk of lung cancer (OR 1.15, 95% CI 1.07-1.24, *P* = 1.72 × 10^-4^). This association was stronger for current and former smokers (OR 1.15, 95% CI 1.09-1.23, *P* = 2.29 × 10^-6^) compared to never smokers (OR 1.09, 95% CI 0.91-1.32, *P* = 0.34). The magnitude of effect was similar across histology types. In analyses of 8,489 cases and 374,506 controls, each copy of the minor allele of rs34536443 was associated with an increased risk of non-Hodgkin lymphoma (OR 1.18, 95% CI 1.05-1.33, *P* = 5.25 × 10^-3^). In secondary analyses, rs34536443 was weakly associated with risk of advanced prostate cancer, but not associated with other cancers examined.

## Discussion

The efficacy of Deucravactinib in treating plaque psoriasis is attributed to the selective inhibition of TYK2, a downstream mediator of pro-inflammatory signalling pathways critical to psoriasis^3^. Using an established partial loss-of-function variant to mimic therapeutic TYK2 inhibition, we show that potential protection from autoimmunity mediated by TYK2 inhibition may be counteracted by an increased risk of lung cancer and non-Hodgkin lymphoma. This finding, including the restriction of an increased lung cancer risk to current and former smokers, is consistent with recent reports of higher rates of these two cancers in users of the JAK inhibitor Tofacitinib in the ORAL Surveillance safety trial^1^. Importantly, the per-allele estimates presented in this analysis may underestimate the effect of therapeutic TYK2 inhibition on cancer given larger TYK2 reductions achieved by Deucravacitinib at doses shown to confer therapeutic benefit (50-80% TYK2 inhibition) compared with the <40% expected per copy of the rs34536443 minor allele^18^. These findings, suggesting potential adverse target-mediated effects of TYK2 inhibition on lung cancer and non-Hodgkin lymphoma, could have important implications for future safety assessment of Deucravacitinib and other TYK2 inhibitors in development.

## Data Availability

Summary genetic association data for cancer endpoints were obtained from the INTEGRAL consortium (https://ilcco.iarc.fr/), PRACTICAL consortium (http://practical.icr.ac.uk/blog/), and GECCO consortium (https://www.fredhutch.org/en/research/divisions/public-health-sciences-division/research/cancer-prevention/genetics-epidemiology-colorectal-cancer-consortium-gecco.html) via approved data usage proposals. Summary genetic association data on breast cancer risk can be downloaded from the Breast Cancer Association Consortium (https://bcac.ccge.medschl.cam.ac.uk/). Summary genetic association data on lymphoma from InterLymph were obtained via dbGaP accession number 15258. Summary genetic association data for the following autoimmune disease analyses were obtained from the GWAS Catalogue (https://www.ebi.ac.uk/gwas/): Crohn's Disease (Study accession: GCST003044), Rheumatoid arthritis (Study accession: GCST002318), Psoriasis (Study accession: GCST005527). Summary genetic association data on Multiple Sclerosis was obtained from the IEU GWAS Catalogue (https://gwas.mrcieu.ac.uk/datasets/ieu-b-18/).

## Acknowledgements

The authors would like to thank the participants of the individual studies contributing to the BCAC, GECCO, CORECT, CCFR, IIBDGC, IMSGC, INTEGRAL-ILCCO, InterLymph, PRACTICAL, and UK Biobank studies. The authors would also like to acknowledge the investigators of these consortia and studies for generating the data used for this analysis. The authors would like to acknowledge the following investigators of the OncoArray and GAME-ON1KG INTEGRAL-ILCCO analyses: Maria Teresa Landi, Victoria Stevens, Ying Wang, Demetrios Albanes, Neil Caporaso, Paul Brennan, Christopher I Amos, Sanjay Shete, Rayjean J Hung, Heike Bickeböller, Angela Risch, Richard Houlston, Stephen Lam, Adonina Tardon, Chu Chen, Stig E Bojesen, Mattias Johansson, H-Erich Wichmann, David Christiani, Gadi Rennert, Susanne Arnold, John K. Field, Loic Le Marchand, Olle Melander, Hans Brunnström, Geoffrey Liu, Angeline Andrew, Lambertus A Kiemeney, Hongbing Shen, Shan Zienolddiny, Kjell Grankvist, Mikael Johansson, M Dawn Teare, Yun-Chul Hong, Jian-Min Yuan, Philip Lazarus, Matthew B Schabath, Melinda C Aldrich. Non-Hodgkin lymphoma data (phs000801) was accessed using dbGaP accession number 15258.

