## Supplementary Material for "Germline TYK2 mutation and cancer risk"

**Non-Hodgkin lymphoma data**

Summary genetic association data for non-Hodgkin lymphoma (NHL) were generated by meta-analysing GWAS data for rs34536443 from two sources: UK Biobank and InterLymph.

Analyses in UK Biobank data used the following ICD9 or ICD10 codes to define NHL cases: 200, 202.0, 202.1, 202.2, 202.7, C82, C83, C84, C85, C86, C88.0, C88.4. Controls were defined as individuals who did not have any cancer code (ICD9 or ICD10) and who did not self-report a cancer diagnosis. GWAS was then conducted using a linear mixed model (LMM) association method as implemented in BOLT-LMM (v2.3) to account for relatedness and population substructure (Loh et al. Nature Genetics. 2015. 47: 284–90). Models were adjusted for sex and genotype array. BOLT-LMM association statistics on the linear scale were then converted to log odds ratios using a Taylor transformation expansion series (Loh et al. Nature Genetics. 2018. 50:906–908). The final model had 2,268 cases and 372,016 controls.

For analyses using InterLymph data, subsequent to variant quality control Chromosome 19 was extracted and exported as VCF. Haplotype phasing was conducted using Eagle (V2.4.1) using the 1000 Genomes phase three data with the parameter --allowRefAltSwap. Minimac4 (V1.0.2) was used to perform imputation using the 1000 Genomes mixed phase three data. The imputed VCF was imported into PLINK 2.0 where SNP 19:10463118:G:C was extracted and the glm parameter selected (no-firth) to perform logistic regression with the following covariates: age, sex, and genetically-derived population principal components. The final model had 6,221 cases and 2,490 controls. Non-Hodgkin lymphoma data (phs000801) was accessed using dbGaP accession number 15258.

Summary statistics obtained from UK Biobank and InterLymph analyses were then meta-analysed using an inverse variance-weighted fixed effects model in METAL (Willer et al. Bioinformatics. 2010. 26: 2190–2191).

**Exploratory smoking initiation analyses**

Given the restriction of an association of the rs34536443 minor allele with lung cancer risk to current and former smokers we examined whether this variant was associated with smoking initiation. To perform this analysis we obtained summary genetic association data on this trait (defined as whether an individual had ever smoked regularly) from a GWAS of 1,232,091 individuals (Liu et al. Nature Genetics. 2019. 51:237–244). There was little evidence of an association of the minor allele of this variant with smoking initiation (unit increase in log odds of ever smoking regularly: 0.0036, 95% CI -0.0131 to 0.0203, *P*=0.67).

**The Colon Cancer Family Registry (CCFR), Colorectal Cancer Transdisciplinary study (CORECT), Genetics and Epidemiology of Colorectal Cancer Consortium (GECCO).**

Sun Ha Jee Jee^1^, Keum Ji Jung Jung^2^, Goncalo R Abecasis Abecasis^3^, Elom K Aglago^4^, Antonio Agudo Agudo^5^, Yoon-Ok Ahn Ahn^6^, Demetrius Albanes (PI) Albanes^7^, M Henar Alonso Alonso^8^, Elizabeth Alwers Alwers^9^, Efrat L Amitay Amitay^10^, Kristin Anderson Anderson^11^, Coral Arnau-Collell Arnau-Collell^12^, Volker Arndt Arndt^13^, Christina Bamia Bamia^14^, Barbara L Banbury Banbury^15^, Richard Barfield Barfield^15^, Elizabeth L Barry Barry^16^, Michael C Bassik Bassik^17^, James W Baurley Baurley^18^, Sonja I Berndt Berndt^7^, Stéphane Bézieau Bézieau^19^, Stephanie A Bien Bien^15^, D Timothy Bishop (PI) Bishop^20^, Juergen Boehm Boehm^21^, Heiner Boeing Boeing^22^, Ivan Borozan Borozan^23^, Emmanouil Bouras Bouras^24^, Marie-Christine Boutron-Ruault Boutron^25^, Hermann Brenner (PI) Brenner^13^, Stefanie Brezina Brezina^26^, Stephan Buch Buch^27^, Daniel D Buchanan Buchanan^28^, Arif Budiarto Budiarto^29^, Susan Bullman Bullman^30^, Andrea Burnett-Hartman Burnett-Hartman^31^, Katja Butterbach Butterbach^32^, Bette J Caan Caan^33^, Qiuyin Cai Cai^34^, Peter T Campbell (PI) Campbell^35^, Federico Canzian Canzian^36^, Yin Cao Cao^37^, Christopher S Carlson Carlson^15^, Prudence Carr Carr^38^, Robert Carreras-Torres Carreras-Torres^39^, Graham Casey Casey^40^, Jose E Castelao Castelao^41^, Antoni Castells Castells^12^, Sergi Castellví-Bel Castellví-Bel^12^, Tjeng Wawan Cenggoro Cenggoro^29^, Andrew T Chan Chan^42^, Jenny Chang-Claude (PI) Chang-Claude^43^, Stephen J Chanock Chanock^7^, Xuechen Chen Chen^44^, Sai Chen Chen^3^, Maria-Dolores Chirlaque Chirlaque^45^, Sang Hee Cho Cho^46^, James Church Church^47^, Gerhard Coetzee Coetzee^48^, Charles Connolly Connolly^15^, David V Conti Conti^49^, Douglas C Corley Corley^50^, Michelle Cotterchio Cotterchio^51^, Chiara Cremolini Cremolini^44^, Amanda J Cross Cross^52^, Marcia Cruz-Correa Cruz Correa^53^, Katarina Cuk Cuk^13^, Keith R Curtis Curtis^15^, James Dai Dai^15^, Mauro D’Amato D'Amato^54^, Christopher H Dampier Dampier^55^, Albert de la Chapelle (deceased) de la Chapelle^56^, Matthew Devall Devall^57^, Brenda Diergaarde Diergaarde^58^, Virginia Diez-Obrero Diez Obrero^59^, Niki Dimou Dimou^60^, Kimberly F Doheny Doheny^61^, David Drew Drew^62^, Mulong Du Du^63^, Margaret Du Du^64^, David Duggan Duggan^65^, Douglas F Easton Easton^66^, Chistopher K Edlund Edlund^49^, Sjoerd G Elias Elias^67^, Faye Elliott Elliott^20^, Dallas R English English^68^, Alfredo Falcone Falcone^69^, Edith JM Feskens Feskens^70^, Jane C Figueiredo (PI) Figueiredo^71^, Rocky Fischer Fischer^72^, Liesel M FitzGerald FitzGerald^73^, David Forman Forman^74^, Amy J French French^75^, Charles Fuchs Fuchs^76^, Manuela-Gago-Dominquez Gago^77^, Manish Gala Gala^42^, Steven Gallinger (PI) Gallinger^78^, W. James Gauderman Gauderman^79^, Marios Giannakis Giannakis^80^, Graham G Giles (PI) Giles^73^, Elizabeth Gillanders Gillanders^81^, Edward Giovannucci Giovannucci^82^, Jian Gong Gong^15^, Phyllis J Goodman Goodman^83^, William M Grady Grady^84^, Peyton Greenside Greenside^85^, Joel Greenson Greenson^86^, John S Grove Grove^87^, Stephen B Gruber (PI) Gruber^88^, Andrea Gsur (PI) Gsur^26^, Mark A Guinter Guinter^89^, Marc J Gunter (PI) Gunter^90^, Feng Guo Guo^13^, Robert W Haile Haile^91^, Christopher A. Haiman Haiman^92^, Jochen Hampe Hampe^27^, Heather Hampel Hampel^93^, Sophia Harlid Harlid^94^, Tabitha A Harrison Harrison^15^, Elizabeth Hauser Hauser^95^, Richard B Hayes Hayes^96^, Chad He He^15^, Volker Heinemann Heinemann^97^, Akihisa Hidaka Hidaka^15^, Eric Ji-Hoon-Joo Hi-Hoon-Joo^98^, Philipp Hofer Hofer^26^, Michael Hoffmeister (PI) Hoffmeister^13^, Andreana Natalie Holowatyj Holowatyj^99^, John L Hopper Hopper^68^, Li Hsu Hsu^15^, Wan-Ling Hsu Hsu^100^, Xinwei Hua Hua^15^, Wen-Yi Huang Huang^7^, Yuhan Huang Huang^15^, Thomas J Hudson Hudson^101^, Meredith Hullar Hullar^15^, David J Hunter Hunter^102^, Jeroen R Huyghe Huyghe^15^, Jae Hwan Oh Hwan Oh^6^, Gemma Ibáñez-Sanz Ibañez-Sanz^8^, Gregory E Idos Idos^49^, Liher Imaz Imaz^103^, Roxann Ingersoll Ingersoll^61^, Rebecca Jackson Jackson^104^, Eric J Jacobs Jacobs^89^, Mazda Jenab Jenab^90^, Mark A Jenkins (PI) Jenkins^68^, Jihyoun Jeon Jeon^105^, Wei-Hua Jia Jia^106^, Kristina Jordahl Jordahl^15^, Amit D Joshi Joshi^102^, Corinne E Joshu Joshu^107^, Paul Limburg Limburg^44^, Yoichiro Kamatani Kamatani^108^, Ellen Kampman Kampman^70^, Hyun Min Kang Kang^3^,; Katie Houlahan^44^, Eric kawaguchi Kawaguchi^109^, Temitope O Keku (PI) Keku^110^, Timothy J Key Key^111^, Hyeong Rok Kim Kim^112^, Dong-Hyun Kim Kim^113^, Jeongseon Kim Kim^114^, Andre Kim Kim^79^, Laurence N Kolonel Kolonel^115^, Charles Kooperberg Kooperberg^15^, Tilman Kuhn Kuhn^116^, Anshul Kundaje Kundaje^17^, Sébastien Küry Küry^19^, Sun-Seog Kweon Kweon^117^, Carlo La Vecchia La Vecchia^118^, Julia D Labadie Labadie^15^, Iris Landorp Vogelaar Lansdorp Vogelaar^119^, Susanna C Larsson Larsson^120^, Cecelia A Laurie Laurie^121^, Loic Le Marchand (PI) Le Marchand^122^, Suzanne M Leal Leal^123^, Soo Chin Lee Lee^124^, Jeffrey K Lee Lee^125^, Flavio Lejbkowicz Lejbkowicz^126^, Mathieu Lemire Lemire^127^, Heinz-Josef Lenz Lenz^128^, David M Levine Levine^121^, Juan Pablo Lewinger Lewinger^129^, Christopher I Li Li^15^, Li Li (PI) Li^130^, Wolfgang Lieb Lieb^131^, Yi Lin Lin^15^, Annika Lindblom Lindblom^132^, Noralane M Lindor Lindor^133^, Sara Lindstroem Lindstroem^15^, Hua Ling Ling^61^, Yun-Ru Liu Liu^134^, Jirong Long Long^34^, Tin L Louie Louie^100^, Fotios Loupakis Loupakis^135^, Yingchang Lu Lu^34^, Frank Luh Luh^136^, Wenjie Ma Ma^137^, Bharuno Mahesworo Mahesworo^18^, Satu Männistö Männistö^138^, Elaine Mardis Mardis^139^, Sanford D Markowitz Markowitz^140^, Vicente Martín Martín^45^, Marie Elena Martinez Martinez^141^, Giovanna Masala Masala^142^, Koichi Matsuda Matsuda^143^, Keitaro Matsuo Matsuo^144^, Kevin J McDonnell McDonnell^145^, Caroline E McNeil McNeil^146^, Leah Mechanic Mechanic^81^, Robert Meester Meester^44^, Marilena Melas Melas^49^, Cameron Miller Miller^147^, Roger L Milne Milne^73^, Jessica Minnier Minnier^148^, Mereia Obon-Santacana Obon-Santacana^149^, Leticia Moreira Moreira^12^, Victor Moreno (PI) Moreno^150^, Lorena Moreno Moreno^12^, John Morrison Morrison^151^, Bhramar Mukherjee Mukherjee^152^, Victor Muñoz-Garzón Muñoz-Garzón^44^, Neil Murphy Murphy^90^, Robin Myte Myte^94^, Alessio Naccarati Naccarati^153^, Hongmei Nan Nan^154^, Rami Nassir Nassir^155^, Ferran Moratalla Navarro Navarro^156^, Sarah C Nelson Nelson^121^, Polly A Newcomb (PI) Newcomb^15^, Christina Newton Newton^157^, Nick Mancuso Mancuso^44^, Deborah A Nickerson Nickerson^158^, Reiko Nishihara Nishihara^159^, Johnathan A Nowak Nowak^159^, Kenneth Offit (PI) Offit^160^, Shuji Ogino (PI) Ogino^161^, N Charlotte Onland-Moret Onland-Moret^67^, Jennifer Ose Ose^162^, Isao Oze Oze^144^, Rish K Pai (PI) Pai^163^, Julie R Palmer (PI) Palmer^164^, Mala Pande Pande^165^, Salvatore Panico Panico^166^, Nick Papadimitrou Papadimitrou^60^, Nicholas Papadopolous Papadopolous^167^, Bens Pardamean Pardamean^18^, Barbara Pardini Pardini^168^, Patrick S Parfrey Parfrey^169^, Rachel Pearlman Pearlman^93^, Anita Peoples Peoples^170^, Vittorio Perduca Perduca^171^, Aurora Perez-Cornago Perez-Cornago^111^, Julyann Pérez-Mayoral Pérez-Mayoral^172^, Ulrike Peters Peters^15^, Elleke Peterse Peterse^119^, Paneen Petersen Petersen^173^, Paul D P Pharoah (PI) Pharoah^66^, Amanda I Phipps Phipps^174^, Mila Pinchev Pinchev^175^, Elizabeth A Platz (PI) Platz^107^, Sarah Plummer Plummer^176^, John D Potter Potter^15^, Ross L Prentice Prentice^15^, Elizabeth Pugh Pugh^61^, Lihong Qi Qi^177^, Qing Zhang Qing^178^, Xuejun Qin Qing^179^, Conghui Qu Qu^15^, Chenxu Qu Qu^180^, Leon Raskin Raskin^181^, Gad Rennert Rennert^182^, Hedy S Rennert Rennert^182^, Elio Riboli Riboli^183^, Miguel Rodríguez-Barranco Rodríguez-Barranco^184^, Joshua Smith Roth^185^, Edward Ruiz-Narvaez Ruiz-Narvaez^186^, Lori C Sakoda (PI) Sakoda^50^, Robert S Sandler (PI) Sandler^110^, Peter C Scacheri Scacheri^187^, Clemens Schafmayer Schafmayer^188^, Stephanie L Schmit Schmit^189^, Jennifer L Schneider Schneider^50^, Robert E Schoen (PI) Schoen^190^, Fredrick R Schumacher Schumacher^191^, Daniela Seminara Seminara^81^, Gianluca Severi Severi^192^, Mitul Shah Shah^193^, Anna Shcherbina Shcherbina^17^, Tameka Shelford Shelford^61^, David Shibata Shibata^194^, Min-Ho Shin Shin^117^, Aesun Shin Shin^195^, Xiao-Ou Shu Shu^196^, Katerina Shulman Shulman^197^, Erin Siegel Siegel^198^, Sabina Sieri Sieri^199^, Nasa A Sinnott-Armstrong Sinnott-Armstrong^17^, Martha L Slattery (PI) Slattery^200^, Joshua D Smith Smith^158^, Linda Snetselaar Snetselaar^201^, Mingyang Song Song^202^, Melissa C Southey Southey^203^, Zsofia K Stadler Stadler^204^, Christa Stegmaier Stegmaier^205^, Robert S Steinfelder Steinfelder^15^, Mariana C Stern Stern^49^, Sebastian Stintzing Stintzing^206^, Yu-Ru Su Su^15^, Wei Sun Sun^15^, Malin Sund Sund^207^, Catherine M Tangen (PI) Tangen^83^, Evi Theodoratou Theodoratou^44^, Stephen N Thibodeau Thibodeau^208^, Duncan C Thomas Thomas^49^, Sushma S Thomas Thomas^15^, Minta Thomas Thomas^15^, Yu Tian Tian^209^, Amanda E Toland Toland^210^, Antonia Trichopoulou Trichopoulou^14^, Quang M Trinh Trinh^211^, Kostas Tsilidis Tsilidis^212^, Natalia Udaltsova Udaltsova^50^, Tomotaka Ugai Ugai^213^, Cornelia M Ulrich Ulrich^21^, Caroline Y Um Um^89^, David J Van Den Berg Van Den Berg^49^, Franzel JB van Duijnhoven van Duijnhoven^70^, Bethany Van Guelpen (PI) Van Guelpen^94^, Henk van Kranen van Kranen^214^, Joseph Vijai Vijai^204^, Paolo Vineis Vineis^215^, Kala Visvanathan (PI) Visvanathan^107^, Pavel Vodicka (PI) Vodicka^153^, Ludmila Vodickova Vodickova^153^, Veronika Vymetalkova Vymetalkova^153^, Michael Wainberg Wainberg^216^, Hansong Wang Wang^122^, Jun Wang Wang^217^, Meilin Wang^63^, Xiaoliang Wang Wang^15^, Ching-Yun Wang^218^, Korbinian Weigl Weigl^13^, Stephanie J Weinstein Weinstein^7^, Vera Wesselink Wesselink^44^, Emily White (PI) White^15^, Lynne R. Wilkens Wilkens^219^, Aung Ko Win Win^68^, C Roland Wolf Wolf^220^, Alicja Wolk (PI) Wolk^120^, Michael O Woods (PI) Woods^221^, Anna H Wu Wu^222^, Yong-Bing Xiang Xiang^223^, Yen Yun Yen^224^,; Yunqi Li^225^, Syed H Zaidi Zaidi^101^, Brent W Zanke Zanke^226^, Ann G Zauber Zauber^227^, Natalia Zemlianskaia Zemlianskaia^129^, Yi-Xin Zeng Zeng^106^, Xuehong Zhang Zhang^228^, Wei Zheng Zheng^229^, Jiayin Zheng Zheng^230^, Yingye Zheng Zheng^231^

^1^Department of Epidemiology and Health Promotion, Graduate School of Public Health, Yonsei University, Seoul, Korea., ^2^Institute for Health Promotion, Graduate School of Public Health, Yonsei University, Seoul, Korea., ^3^Department of Biostatistics and Center for Statistical Genetics, University of Michigan, Ann Arbor, Michigan, USA., ^4^International Agency for Research on Cancer (IARC-WHO), Lyon, France., ^5^Unit of Nutrition and Cancer, Cancer Epidemiology Research Program, Catalan Institute of Oncology-IDIBELL, L'Hospitalet de Llobregat, Barcelona, Spain., ^6^Department of Preventive Medicine, Seoul National University College of Medicine, Seoul, South Korea., ^7^Division of Cancer Epidemiology and Genetics, National Cancer Institute, National Institutes of Health, Bethesda, Maryland, USA., ^8^Cancer Prevention and Control Program, Catalan Institute of Oncology-IDIBELL, L'Hospitalet de Llobregat, Barcelona, Spain., ^9^Division of Clinical Epidemiology and Aging Research , German Cancer Research Center , Heidelberg , Germany, ^10^Division of Clinical Epidemiology and Aging Research, German Cancer Research Center, Heidelberg, Germany., ^11^Division of Epidemiology and Community Health, University of Minnesota, Minneapolis, Minnesota, USA., ^12^Gastroenterology Department, Hospital Clínic, Institut d'Investigacions Biomèdiques August Pi i Sunyer (IDIBAPS), Centro de Investigación Biomédica en Red de Enfermedades Hepáticas y Digestivas (CIBEREHD), University of Barcelona, Barcelona, Spain., ^13^Division of Clinical Epidemiology and Aging Research, German Cancer Research Center (DKFZ), Heidelberg, Germany., ^14^Hellenic Health Foundation, Athens, Greece., ^15^Public Health Sciences Division, Fred Hutchinson Cancer Research Center, Seattle, Washington, USA., ^16^Department of Epidemiology, Geisel School of Medicine at Dartmouth, Hanover, NH, USA., ^17^Department of Genetics, Stanford University, Stanford, California, USA., ^18^Bioinformatics and Data Science Research Center, Bina Nusantara University, Jakarta, Indonesia., ^19^Service de Génétique Médicale, Centre Hospitalier Universitaire (CHU) Nantes, Nantes, France., ^20^Leeds Institute of Cancer and Pathology, University of Leeds, Leeds, UK., ^21^Huntsman Cancer Institute and Department of Population Health Sciences, University of Utah, Salt Lake City, Utah, USA., ^22^Department of Epidemiology, German Institute of Human Nutrition (DIfE), Potsdam-Rehbrücke, Germany., ^23^Ontario Institute for Cancer Research, Toronto, Canada., ^24^Laboratory of Hygiene, Social & Preventive Medicine and Medical Statistics, Department of Medicine, School of Health Sciences, Aristotle University of Thessaloniki, Greece., ^25^Inserm U1018, Center for Research in Epidemiology and Population Health (CESP), Gustave Roussy, Villejuif, France., ^26^Institute of Cancer Research, Department of Medicine I, Medical University Vienna, Vienna, Austria., ^27^Department of Medicine I, University Hospital Dresden, Technische Universität Dresden (TU Dresden), Dresden, Germany., ^28^Colorectal Oncogenomics Group, Department of Clinical Pathology, The University of Melbourne, Parkville, Victoria 3010 Australia, ^29^Bioinformatics and Data Science Research Center, Bina Nusantara University, Jakarta, Indonesia, ^30^Human Biology Division, Fred Hutchinson Cancer Research Center, Seattle, Washington, USA., ^31^Institute for Health Research, Kaiser Permanente Colorado, Denver, Colorado, USA., ^32^Division of Clinical Epidemiology and Aging Research, German Cancer Research Center (DKFZ), Heidelberg, Germany.; Unit of Biomarkers & Susceptibility, Oncology Data Analytics Program, Catalan Institute of Oncology (ICO), 08908 Hospitalet de Llobregat, Barcelona, Spain.; Biomedical Research Centre Network for Epidemiology & Public Health (CIBERESP), 28029 Madrid, Spain., ^33^Division of Research, Kaiser Permanente Medical Care Program, Oakland, California, USA., ^34^Division of Epidemiology, Department of Medicine, Vanderbilt-Ingram Cancer Center, Vanderbilt Epidemiology Center, Vanderbilt University School of Medicine, Nashville Tennessee, USA., ^35^Department of Epidemiology and Population Health, Albert Einstein College of Medicine, Bronx, NY, USA., ^36^Genomic Epidemiology Group, German Cancer Research Center (DKFZ), Heidelberg, Germany., ^37^Division of Public Health Sciences, Department of Surgery, Washington University School of Medicine, St Louis, Missouri, USA., ^38^Division of Clinical Epidemiology, German Cancer Research Center, Heidelberg, Germany., ^39^Colorectal Cancer Group, ONCOBELL Program, Bellvitge Biomedical Research Institute (IDIBELL), L'Hospitalet de Llobregat, 8908 Barcelona, Spain., ^40^Center for Public Health Genomics, University of Virginia, Charlottesville, Virginia, USA., ^41^Instituto de Investigación Sanitaria Galicia Sur (IISGS), Xerencia de Xestion Integrada de Vigo-SERGAS, Oncology and Genetics Unit, Vigo, Spain., ^42^Division of Gastroenterology, Massachusetts General Hospital and Harvard Medical School, Boston, Massachusetts, USA., ^43^Division of Cancer Epidemiology, German Cancer Research Center (DKFZ), Heidelberg, Germany., ^44^(No affiliation data provided), ^45^CIBER Epidemiología y Salud Pública (CIBERESP), Madrid, Spain., ^46^Department of Hematology-Oncology, Chonnam National University Hospital, Hwasun, South Korea., ^47^Department of Colorectal Surgery, Cleveland Clinic, Cleveland, Ohio 44195, USA., ^48^Van Andel Research Institute, Grand Rapids, Michigan 49502, USA., ^49^Department of Population and Public Health Sciences, Keck School of Medicine, University of Southern California, Los Angeles, California, USA., ^50^Division of Research, Kaiser Permanente Northern California, Oakland, California, USA., ^51^Prevention and Cancer Control, Cancer Care Ontario, Toronto, ON, Canada., ^52^Department of Epidemiology and Biostatistics, Imperial College London, London, UK., ^53^Comprehensive Cancer Center, University of Puerto Rico Medical Sciences Campus, San Juan, PR 00936, USA., ^54^CIC bioGUNE - BRTA, Bizkaia Science and Technology Park, Spain, ^55^Department of General Surgery, University of Virginia School of Medicine, Charlottesville, Virginia, USA., ^56^Department of Cancer Biology and Genetics and the Comprehensive Cancer Center, The Ohio State University, Columbus, Ohio, USA., ^57^Center for Public Health Genomics, Department of Public Health Sciences, University of Virginia, Charlottesville, Virginia, USA, ^58^Department of Human Genetics, Graduate School of Public Health, University of Pittsburgh, Pittsburgh, Pennsylvania, USA., ^59^Unit of Biomarkers and Susceptibility, Oncology Data Analytics Program, Catalan Institute of Oncology, Barcelona 08908, Spain., ^60^Section of Nutrition and Metabolism, International Agency for Research on Cancer, Lyon, France., ^61^Center for Inherited Disease Research (CIDR), Institute of Genetic Medicine, Johns Hopkins University School of Medicine, Baltimore, Maryland, USA., ^62^Clinical & Translational Epidemiology Unit, Massachusetts General Hospital and Harvard Medical School, Boston, MA, USA., ^63^School of Public Health, Nanjing Medical University, ^64^Department of Epidemiology and Biostatistics, Memorial Sloan Kettering Cancer Center, New York, NY, USA., ^65^Translational Genomics Research Institute - An Affiliate of City of Hope, Phoenix, Arizona, USA., ^66^Department of Public Health and Primary Care, University of Cambridge, Cambridge, UK., ^67^Julius Center for Health Sciences and Primary Care, University Medical Center Utrecht, Utrecht, The Netherlands., ^68^Centre for Epidemiology and Biostatistics, Melbourne School of Population and Global Health, The University of Melbourne, Melbourne, Victoria, Australia., ^69^;, ^70^Division of Human Nutrition, Wageningen University and Research, Wageningen, The Netherlands., ^71^Department of Medicine, Samuel Oschin Comprehensive Cancer Institute, Cedars-Sinai Medical Center, Los Angeles, CA, USA., ^72^University of Michigan Comprehensive Cancer Center, Ann Arbor, Michigan, USA., ^73^Cancer Epidemiology Division, Cancer Council Victoria, Melbourne, Victoria, Australia., ^74^International Agency for Research on Cancer, World Health Organization, Lyon, France., ^75^Division of Laboratory Genetics, Department of Laboratory Medicine and Pathology, Mayo Clinic, 200 First Street SW, Rochester, MN, 55905, USA., ^76^Department of Medical Oncology, Dana-Farber Cancer Institute, Brookline, Massachusetts 02115, USA., ^77^Fundación Gallega de Medicina Genómica, Grupo de Genética del Cáncer, Instituto de Investigación Sanitaria de Santiago IDIS, Complejo Hospitalario Univ. Santiago-CHUS, SERGAS, Spain, ^78^Lunenfeld Tanenbaum Research Institute, Mount Sinai Hospital, University of Toronto, Toronto, Ontario, Canada., ^79^Division of Biostatistics, Department of Population and Public Health Sciences, Keck School of Medicine, University of Southern California, Los Angeles, California, USA., ^80^Department of Medical Oncology, Dana-Farber Cancer Institute, Boston, Massachusetts, USA, ^81^Division of Cancer Control and Population Sciences, National Cancer Institute, Bethesda, Maryland, USA., ^82^Harvard Medical School, Boston, Massachusetts 02114, USA., ^83^SWOG Statistical Center, Fred Hutchinson Cancer Research Center, Seattle, Washington, USA., ^84^Clinical Research Division, Fred Hutchinson Cancer Research Center, Seattle, Washington, USA., ^85^Department of Biomedical Data Science, Stanford University, Stanford, California, USA., ^86^Department of Pathology, University of Michigan, Ann Arbor, Michigan 48104, USA., ^87^University of Hawaii Cancer Research Center, Honolulu, Hawaii, USA., ^88^Department of Preventive Medicine & USC Norris Comprehensive Cancer Center, Keck School of Medicine, University of Southern California, Los Angeles, California, USA., ^89^Behavioral and Epidemiology Research Group, American Cancer Society, Atlanta, Georgia, USA., ^90^Nutrition and Metabolism Section, International Agency for Research on Cancer, World Health Organization, Lyon, France., ^91^Samuel Oschin Comprehensive Cancer Institute, CEDARS-SINAI, Los Angeles, California, USA., ^92^Center for Genetic Epidemiology, Department of Population and Public Health Sciences, Keck School of Medicine, University of Southern California, Los Angeles, California, USA., ^93^Division of Human Genetics, Department of Internal Medicine, The Ohio State University Comprehensive Cancer Center, Columbus, Ohio, USA., ^94^Department of Radiation Sciences, Oncology Unit, Umeå University, Umeå, Sweden., ^95^VA Cooperative Studies Program Epidemiology Center, Durham Veterans Affairs Health Care System, Durham, North Carolina, USA., ^96^Division of Epidemiology, Department of Population Health, New York University School of Medicine, New York, New York, USA., ^97^Department of Medical Oncology and Comprehensive Cancer Center, Ludwig Maximilians University - Grosshadern, Munich, Germany., ^98^Department of Paediatrics; University of Melbourne; Parkville, VIC Australia; Cancer, Disease and Developmental Epigenetics Group; Murdoch Childrens Research Institute (MCRI); Royal Children's Hospital; Parkville, VIC Australia., ^99^Huntsman Cancer Institute, Salt Lake City, UT; Department of Population Health Sciences, University of Utah, Salt Lake City, UT; Department of Medicine, Vanderbilt University Medical Center, Nashville, TN; Vanderbilt-Ingram Cancer Center, Nashville, TN. Electronic address:., ^100^University of Washington, Department of Biostatistics, Seattle, Washington, USA., ^101^Ontario Institute for Cancer Research, Toronto, Ontario, Canada., ^102^Department of Epidemiology, Harvard T.H. Chan School of Public Health, Harvard University, Boston, Massachusetts, USA., ^103^Public Health Division of Gipuzkoa, Health Department of Basque Country, Spain., ^104^Department of Internal Medicine, Ohio State Medical Center, Columbus, Ohio, USA., ^105^Department of Epidemiology, University of Michigan, Ann Arbor, Michigan, USA., ^106^State Key Laboratory of Oncology in South China, Cancer Center, Sun Yat-sen University, Guangzhou, China., ^107^Department of Epidemiology, Johns Hopkins Bloomberg School of Public Health, Baltimore, Maryland, USA., ^108^Laboratory for Statistical Analysis, RIKEN Center for Integrative Medical Sciences, Kanagawa, Japan., ^109^Department of Biostatistics, Fielding School of Public Health, University of California, Los Angeles, Los Angeles, CA, USA., ^110^Center for Gastrointestinal Biology and Disease, University of North Carolina, Chapel Hill, North Carolina, USA., ^111^Cancer Epidemiology Unit, Nuffield Department of Population Health, University of Oxford, Oxford, UK., ^112^Department of Surgery, Chonnam National University Hwasun Hospital and Medical School, Hwasun, Korea., ^113^Department of Social and Preventive Medicine, Hallym University College of Medicine, Okcheon-dong, South Korea, ^114^Department of Cancer Biomedical Science, Graduate School of Cancer Science and Policy, National Cancer Center, Gyeonggi-do, South Korea., ^115^Office of Public Health Studies, University of Hawaii Manoa, Honolulu, Hawaii, USA., ^116^Division of Cancer Epidemiology German Cancer Research Center (DKFZ) Heidelberg, Germany., ^117^Department of Preventive Medicine, Chonnam National University Medical School, Gwangju, Korea., ^118^Hellenic Health Foundation,Athens, Greece; Department of Clinical Sciences and Community Health, Università degli Studi di Milano, Milan, Italy., ^119^Department of Public Health, Erasmus MC, University Medical Center, Rotterdam, The Netherlands., ^120^Institute of Environmental Medicine, Karolinska Institutet, Stockholm, Sweden., ^121^Department of Biostatistics, University of Washington, Seattle, Washington, USA., ^122^University of Hawaii Cancer Center, Honolulu, Hawaii, USA., ^123^Department of Molecular and Human Genetics, Baylor College of Medicine, Houston, Texas, USA., ^124^National University Cancer Institute, Singapore., ^125^Department of Gastroenterology, Kaiser Permanente San Francisco Medical Center, San Francisco, California, USA., ^126^The Clalit Health Services, Personalized Genomic Service, Carmel, Haifa, Israel., ^127^PanCuRx Translational Research Initiative, Ontario, Institute for Cancer Research, Toronto, Ontario, Canada., ^128^Department of Medicine, University of Southern California, Los Angeles, CA, ^129^Department of Preventive Medicine, University of Southern California, Los Angeles, California., ^130^Department of Family Medicine, University of Virginia, Charlottesville, Virginia, USA., ^131^Institute of Epidemiology, PopGen Biobank, Christian-Albrechts-University Kiel, Kiel, Germany., ^132^Department of Clinical Genetics, Karolinska University Hospital, Stockholm, Sweden., ^133^Department of Health Science Research, Mayo Clinic, Scottsdale, Arizona, USA., ^134^Office of Human Research, Taipei Medical University, Taipei, Taiwan., ^135^Unit of Oncology 1 - Department of Oncology, Istituto Oncologico Veneto, IRCCS Padua, Italy., ^136^Sino-American Cancer Foundation, Covina, CA, USA, ^137^Clinical and Translational Epidemiology Unit, Massachusetts General Hospital, MA, USA.; Division of Gastroenterology, Massachusetts General Hospital and Harvard Medical School, Boston, MA, USA., ^138^Department of Public Health Solutions, National Institute for Health and Welfare, Helsinki, Finland., ^139^The Steve and Cindy Rasmussen Institute for Genomic Medicine, Nationwide Children's Hospital, Columbus, Ohio, USA.; Department of Pediatrics, The Ohio State University, Columbus, Ohio, USA., ^140^Departments of Medicine and Genetics, Case Comprehensive Cancer Center, Case Western Reserve University, and University Hospitals of Cleveland, Cleveland, Ohio, USA., ^141^University of California San Diego, Moores Cancer Center, La Jolla, California, USA., ^142^Cancer Risk Factors and Life-Style Epidemiology Unit, Institute of Cancer Research, Prevention and Clinical Network - ISPRO, Florence, Italy., ^143^Laboratory of Clinical Genome Sequencing, Department of Computational Biology and Medical Sciences, Graduate School of Frontier Sciences, University of Tokyo, Tokyo, Japan., ^144^Division of Molecular and Clinical Epidemiology, Aichi Cancer Center Research Institute, Nagoya, Japan., ^145^City of Hope National Medical Center, Duarte, California, USA, ^146^USC Norris Comprehensive Cancer Center, University of Southern California, Los Angeles, California, USA., ^147^Department of Public Health Sciences, Medical University of South Carolina, Charleston, SC., ^148^School of Public Health , Oregon Health & Science University , Portland , OR , USA., ^149^Unit of Nutrition, Environment and Cancer, Cancer Epidemiology Research Program, Catalan Institute of Oncology (ICO-IDIBELL), Avda Gran Via Barcelona 199-203, 08908L'Hospitalet de Llobregat, Barcelona, Spain., ^150^Oncology Data Analytics Program, Catalan Institute of Oncology-IDIBELL, L'Hospitalet de Llobregat, Barcelona, Spain., ^151^Department of Population and Public Health Sciences, Keck School of Medicine of the University of Southern California, Los Angeles, CA, USA.., ^152^University of Michigan Comprehensive Cancer Center, Ann Arbor, Michigan 48105, USA., ^153^Department of Molecular Biology of Cancer, Institute of Experimental Medicine of the Czech Academy of Sciences, Prague, Czech Republic., ^154^Department of Epidemiology, Richard M. Fairbanks School of Public Health , Indianapolis , Indiana , USA., ^155^Department of Pathology, School of Medicine, Umm Al-Qura’a University, Saudi Arabia, ^156^Oncology Data Analytics Program, Catalan Institute of Oncology, L'Hospitalet de Llobregat, Barcelona, Spain; Consortium for Biomedical Research in Epidemiology and Public Health, Madrid, Spain; Department of Clinical Sciences, Faculty of Medicine, University of Barcelona, Barcelona, Spain., ^157^Department of Population Science, American Cancer Society, Atlanta, Georgia., ^158^Department of Genome Sciences, University of Washington, Seattle, Washington, USA., ^159^Program in MPE Molecular Pathological Epidemiology, Department of Pathology, Brigham and Women's Hospital and Harvard Medical School, Boston, Massachusetts, USA, ^160^Clinical Genetics Service, Department of Medicine, Memorial Sloan-Kettering Cancer Center, New York, New York, USA., ^161^Program in MPE Molecular Pathological Epidemiology, Department of Pathology, Brigham and Women's Hospital, Harvard Medical School, Boston, Massachusetts, USA., ^162^Huntsman Cancer Institute, University of Utah, Salt Lake City, Utah.; Department of Population Health Sciences, University of Utah, Salt Lake City, Utah., ^163^Department of Laboratory Medicine and Pathology, Mayo Clinic Arizona, Scottsdale, Arizona, USA., ^164^Boston University School of Medicine, ^165^Department of Gastroenterology, Hepatology and Nutrition, The University of Texas MD Anderson Cancer Center, Houston, TX, 77030, USA., ^166^Dipartimento di Medicina Clinica e Chirurgia, Federico II University, Naples, Italy., ^167^Department of Sarcoma Medical Oncology, The University of Texas M. D. Anderson Cancer Center, Houston, Texas 77030, USA., ^168^Italian Institute for Genomic Medicine (IIGM), Turin, Italy., ^169^Memorial University, Faculty of Medicine, Newfoundland, Canada., ^170^Division of Population Sciences, Huntsman Cancer Institute, Salt Lake City, Utah, USA.; Department of Population Health Sciences, University of Utah, Salt Lake City, UT, USA., ^171^Laboratoire de Mathématiques Appliquées MAP5 (UMR CNRS 8145), Université Paris Descartes, Paris, France., ^172^Division of Cancer Biology, University of Puerto Rico Comprehensive Cancer Center, San Juan, Puerto Rico., ^173^Department of Epidemiology, University of Washington School of Public Health, Seattle, Washington.; Public Health Sciences, Fred Hutchinson Cancer Research Center, Seattle, Washington., ^174^Department of Epidemiology, University of Washington, Seattle, WA, USA., ^175^Department of Community Medicine and Epidemiology, Carmel Medical Center, Haifa, Israel., ^176^Center for Public Health Genomics, University of Virginia, Charlottesville, Virginia 22908, USA., ^177^Department of Public Health Sciences, University of California Davis, Davis, California, USA., ^178^Genomics Shared Resource, Fred Hutchinson Cancer Research Center, Seattle, Washington, USA., ^179^Duke Molecular Physiology Institute, Duke University, Durham, NC, USA, ^180^USC Norris Comprehensive Cancer Center, Keck School of Medicine, University of Southern California, Los Angeles, California, USA., ^181^Division of Epidemiology, Vanderbilt Epidemiology Center, Vanderbilt University School of Medicine, Nashville, Tennessee, USA., ^182^Department of Community Medicine and Epidemiology, Lady Davis Carmel Medical Center, Haifa, Israel., ^183^School of Public Health, Imperial College London, London, UK., ^184^Escuela Andaluza de Salud Pública, Instituto de Investigación Biosanitaria ibs.GRANADA, Hospitales Universitarios de Granada/Universidad de Granada, Granada, Spain., ^185^Department of Genome Sciences, University of Washington, Seattle, WA, United States., ^186^Department of Nutritional Sciences, University of Michigan School of Public Health, Ann Arbor, Michigan, USA., ^187^Department of Genetics and Genome Sciences, Case Western Reserve University, Cleveland, Ohio, USA., ^188^Department of General Surgery, University Hospital Rostock, Rostock, Germany., ^189^Genomic Medicine Institute, Cleveland Clinic, Cleveland, Ohio, USA., ^190^Department of Medicine and Epidemiology, University of Pittsburgh Medical Center, Pittsburgh, Pennsylvania, USA., ^191^Department of Population and Quantitative Health Sciences, Case Western Reserve University, Cleveland, Ohio, USA., ^192^Center Epidemiology Centre, Cancer Council Victoria, Melbourne, Victoria 3004, Australia., ^193^Centre for Cancer Genetic Epidemiology, Department of Oncology, University of Cambridge, Cambridge, UK., ^194^Department of Surgery, University of Tennessee Health Science Center, Memphis, TN, USA., ^195^Department of Preventive Medicine, Seoul National University College of Medicine, Seoul National University Cancer Research Institute, Seoul, South Korea., ^196^Vanderbilt University Medical Center, Nashville Tennessee, USA., ^197^Oncology Unit, Hillel Yaffe Medical Center, Hadera, Israel., ^198^Cancer Epidemiology Program, H. Lee Moffitt Cancer Center and Research Institute, Tampa, FL, USA., ^199^Epidemiology and Prevention Unit, Fondazione IRCCS Istituto Nazionale dei Tumori, Milan, Italy., ^200^Department of Internal Medicine, University of Utah, Salt Lake City, Utah, USA., ^201^Department of Epidemiology, University of Iowa College of Public Health , Iowa City , Iowa , USA., ^202^Clinical and Translational Epidemiology Unit, Massachusetts General Hospital and Harvard Medical School, Boston, Massachusetts, USA., ^203^Precision Medicine, School of Clinical Sciences at Monash Health, Monash University, Clayton, Victoria, Australia., ^204^Department of Medicine, Memorial Sloan Kettering Cancer Center, New York, New York, USA., ^205^Saarland Cancer Registry, Saarbrücken, Germany., ^206^Department of Hematology and Oncology, University of Munich (LMU), Munich, Germany., ^207^Department of Surgical and Perioperative Sciences, Umeå University, Umeå, Sweden., ^208^Division of Laboratory Genetics, Department of Laboratory Medicine and Pathology, Mayo Clinic, Rochester, Minnesota, USA., ^209^Division of Cancer Epidemiology, German Cancer Research Center (DKFZ), Heidelberg, Germany, ^210^Departments of Cancer Biology and Genetics and Internal Medicine, Comprehensive Cancer Center, The Ohio State University, Columbus, Ohio, USA., ^211^Ontario Institute for Cancer Research, 661 University Avenue, Toronto, Ontario, Canada., ^212^Department of Epidemiology and Biostatistics, Imperial College London, School of Public Health, London, UK., ^213^Research Fellow, Harvard T.H. Chan School of Public Health, Department of Epidemiology, Boston, Massachusetts, USA., ^214^National Institute for Public Health and the Environment (RIVM), Bilthoven, The Netherlands., ^215^Centre for Environment and Health School of Public Health Imperial College London St Mary's Campus Norfolk Place, London, United Kingdom., ^216^Department of Computer Science, Stanford University, Stanford, California, USA., ^217^Division of Cancer Epidemiology and Genetics, Cancer Genomics Research Laboratory, National Cancer Institute, Division of Cancer Epidemiology and Genetics, SAIC-Frederick, Inc., Frederick National Laboratory for Cancer Research, Frederick, MD, USA, Department of Biochemistry and Centre for Genomic Sciences, LKS Faculty of Medicine, The University of Hong Kong, Hong Kong SAR, China., ^218^Public Health Sciences Division, Fred Hutchinson Cancer Research Center, Seattle Washington, USA., ^219^University of Hawaii Cancer Centre, Honolulu, HI, USA., ^220^School of Medicine, University of Dundee, Dundee, Scotland., ^221^Memorial University of Newfoundland, Discipline of Genetics, St. John's, Canada., ^222^University of Southern California, Preventative Medicine, Los Angeles, California, USA., ^223^State Key Laboratory of Oncogenes and Related Genes & Department of Epidemiology, Shanghai Cancer Institute, Renji Hospital, Shanghai Jiaotong University School of Medicine, Shanghai, China., ^224^Taipei Medical University, Taipei, Taiwan, ^225^Yunqi Li, ^226^University of Ottawa, Division of Hematology, Ottawa, Canada., ^227^Department of Epidemiology and Biostatistics, Memorial Sloan Kettering Cancer Center, New York, New York, USA., ^228^Channing Division of Network Medicine, Department of Medicine, Brigham and Women's Hospital and Harvard Medical School, Boston, Massachusetts, USA., ^229^Division of Epidemiology, Department of Medicine, Vanderbilt-Ingram Cancer Center, Vanderbilt Epidemiology Center, Vanderbilt University School of Medicine, Nashville, Tennessee, USA., ^230^Division of Public Health Sciences, Fred Hutchinson Cancer Research Center, Seattle, WA, USA., ^231^Division of Public Health Sciences, Fred Hutchinson Cancer Research Center, Seattle, Washington, USA.

**The PRACTICAL Consortium**

[**http://practical.icr.ac.uk/**](http://practical.icr.ac.uk/)

Rosalind A. Eeles^1,2^, Christopher A. Haiman^3^, Zsofia Kote-Jarai^1^, Fredrick R. Schumacher^4,5^, Sara Benlloch^6,1^, Ali Amin Al Olama^6,7^, Kenneth Muir^8,9^, Sonja I. Berndt^10^, David V. Conti^3^, Fredrik Wiklund^11^, Stephen Chanock^10^, Ying Wang^12^, Victoria L. Stevens^12^, Catherine M. Tangen^13^, Jyotsna Batra^14,15^, Judith A. Clements^14,15^, APCB BioResource (Australian Prostate Cancer BioResource)^14,15^, Henrik Grönberg^11^, Nora Pashayan^16,17^, Johanna Schleutker^18,19^, Demetrius Albanes^10^, Stephanie Weinstein^10^, Alicja Wolk^20^, Catharine M. L. West^21^, Lorelei A. Mucci^22^, Géraldine Cancel-Tassin^23,24^, Stella Koutros^10^, Karina Dalsgaard Sørensen^25,26^, Eli Marie Grindedal^27^, David E. Neal^28,29,30^, Freddie C. Hamdy^31,32^, Jenny L. Donovan^33^, Ruth C. Travis^34^, Robert J. Hamilton^35,36^, Sue Ann Ingles^37^, Barry S. Rosenstein^38,39^, Yong-Jie Lu^40^, Graham G. Giles^41,42,43^, Adam S. Kibel^44^, Ana Vega^45,46,47^, Manolis Kogevinas^48,49,50,51^, Kathryn L. Penney^52^, Jong Y. Park^53^, Janet L. Stanford^54,55^, Cezary Cybulski^56^, Børge G. Nordestgaard^57,58^, Sune F. Nielsen^57,58^, Hermann Brenner^59,60,61^, Christiane Maier^62^, Jeri Kim^63^, Esther M. John^64^, Manuel R. Teixeira^65,66,67^, Susan L. Neuhausen^68^, Kim De Ruyck^69^, Azad Razack^70^, Lisa F. Newcomb^54,71^, Davor Lessel^72^, Radka Kaneva^73^, Nawaid Usmani^74,75^, Frank Claessens^76^, Paul A. Townsend^77,78^, Manuela Gago-Dominguez^79,80^, Monique J. Roobol^81^, Florence Menegaux^82^, Kay-Tee Khaw^83^, Lisa Cannon-Albright^84,85^, Hardev Pandha^78^, Stephen N. Thibodeau^86^, David J. Hunter^87^, Peter Kraft^88^, William J. Blot^89,90^, Elio Riboli^91^

^1^The Institute of Cancer Research, London, SM2 5NG, UK
^2^Royal Marsden NHS Foundation Trust, London, SW3 6JJ, UK
^3^Center for Genetic Epidemiology, Department of Preventive Medicine, Keck School of Medicine, University of Southern California/Norris Comprehensive Cancer Center, Los Angeles, CA 90015, USA
^4^Department of Population and Quantitative Health Sciences, Case Western Reserve University, Cleveland, OH 44106-7219, USA
^5^Seidman Cancer Center, University Hospitals, Cleveland, OH 44106, USA.
^6^Centre for Cancer Genetic Epidemiology, Department of Public Health and Primary Care, University of Cambridge, Strangeways Research Laboratory, Cambridge CB1 8RN, UK
^7^University of Cambridge, Department of Clinical Neurosciences, Stroke Research Group, R3, Box 83, Cambridge Biomedical Campus, Cambridge CB2 0QQ, UK
^8^Division of Population Health, Health Services Research and Primary Care, University of Manchester, Oxford Road, Manchester, M13 9PL, UK
^9^Warwick Medical School, University of Warwick, Coventry, CV4 7AL, UK
^10^Division of Cancer Epidemiology and Genetics, National Cancer Institute, NIH, Bethesda, Maryland, 20892, USA
^11^Department of Medical Epidemiology and Biostatistics, Karolinska Institute, SE-171 77 Stockholm, Sweden
^12^Department of Population Science, American Cancer Society, 250 Williams Street, Atlanta, GA 30303, USA
^13^SWOG Statistical Center, Fred Hutchinson Cancer Research Center, Seattle, WA 98109, USA
^14^Australian Prostate Cancer Research Centre-Qld, Institute of Health and Biomedical Innovation and School of Biomedical Sciences, Queensland University of Technology, Brisbane QLD 4059, Australia
^15^Translational Research Institute, Brisbane, Queensland 4102, Australia
^16^Department of Applied Health Research, University College London, London, WC1E 7HB, UK
^17^Centre for Cancer Genetic Epidemiology, Department of Oncology, University of Cambridge, Strangeways Laboratory, Worts Causeway, Cambridge, CB1 8RN, UK
^18^Institute of Biomedicine, University of Turku, Finland
^19^Department of Medical Genetics, Genomics, Laboratory Division, Turku University Hospital, PO Box 52, 20521 Turku, Finland
^20^Department of Surgical Sciences, Uppsala University, 75185 Uppsala, Sweden
^21^Division of Cancer Sciences, University of Manchester, Manchester Academic Health Science Centre, Radiotherapy Related Research, The Christie Hospital NHS Foundation Trust, Manchester, M13 9PL UK
^22^Department of Epidemiology, Harvard T. H. Chan School of Public Health, Boston, MA 02115, USA
^23^CeRePP, Tenon Hospital, F-75020 Paris, France.
^24^Sorbonne Universite, GRC n°5 , AP-HP, Tenon Hospital, 4 rue de la Chine, F-75020 Paris, France
^25^Department of Molecular Medicine, Aarhus University Hospital, Palle Juul-Jensen Boulevard 99, 8200 Aarhus N, Denmark
^26^Department of Clinical Medicine, Aarhus University, DK-8200 Aarhus N
^27^Department of Medical Genetics, Oslo University Hospital, 0424 Oslo, Norway
^28^Nuffield Department of Surgical Sciences, University of Oxford, Room 6603, Level 6, John Radcliffe Hospital, Headley Way, Headington, Oxford, OX3 9DU, UK
^29^University of Cambridge, Department of Oncology, Box 279, Addenbrooke's Hospital, Hills Road, Cambridge CB2 0QQ, UK
^30^Cancer Research UK, Cambridge Research Institute, Li Ka Shing Centre, Cambridge, CB2 0RE, UK
^31^Nuffield Department of Surgical Sciences, University of Oxford, Oxford, OX1 2JD, UK
^32^Faculty of Medical Science, University of Oxford, John Radcliffe Hospital, Oxford, UK
^33^Population Health Sciences, Bristol Medical School, University of Bristol, BS8 2PS, UK
^34^Cancer Epidemiology Unit, Nuffield Department of Population Health, University of Oxford, Oxford, OX3 7LF, UK
^35^Dept. of Surgical Oncology, Princess Margaret Cancer Centre, Toronto ON M5G 2M9, Canada
^36^Dept. of Surgery (Urology), University of Toronto, Canada
^37^Department of Preventive Medicine, Keck School of Medicine, University of Southern California/Norris Comprehensive Cancer Center, Los Angeles, CA 90015, USA
^38^Department of Radiation Oncology and Department of Genetics and Genomic Sciences, Box 1236, Icahn School of Medicine at Mount Sinai, One Gustave L. Levy Place, New York, NY 10029, USA
^39^Department of Genetics and Genomic Sciences, Icahn School of Medicine at Mount Sinai, New York, NY 10029-5674 , USA.
^40^Centre for Cancer Biomarker and Biotherapeutics, Barts Cancer Institute, Queen Mary University of London, John Vane Science Centre, Charterhouse Square, London, EC1M 6BQ, UK
^41^Cancer Epidemiology Division, Cancer Council Victoria, 615 St Kilda Road, Melbourne, VIC 3004, Australia
^42^Centre for Epidemiology and Biostatistics, Melbourne School of Population and Global Health, The University of Melbourne, Grattan Street, Parkville, VIC 3010, Australia
^43^Precision Medicine, School of Clinical Sciences at Monash Health, Monash University, Clayton, Victoria 3168, Australia
^44^Division of Urologic Surgery, Brigham and Womens Hospital, 75 Francis Street, Boston, MA 02115, USA
^45^Fundación Pública Galega Medicina Xenómica, Santiago de Compostela, 15706, Spain.
^46^Instituto de Investigación Sanitaria de Santiago de Compostela, Santiago De Compostela, 15706, Spain.
^47^Centro de Investigación en Red de Enfermedades Raras (CIBERER), Spain
^48^ISGlobal, Barcelona, Spain
^49^IMIM (Hospital del Mar Medical Research Institute), Barcelona, Spain
^50^CIBER Epidemiología y Salud Pública (CIBERESP), 28029 Madrid, Spain
^51^Universitat Pompeu Fabra (UPF), Barcelona, Spain
^52^Channing Division of Network Medicine, Department of Medicine, Brigham and Women's Hospital/Harvard Medical School, Boston, MA 02115, USA
^53^Department of Cancer Epidemiology, Moffitt Cancer Center, 12902 Magnolia Drive, Tampa, FL 33612, USA
^54^Division of Public Health Sciences, Fred Hutchinson Cancer Research Center, Seattle, Washington, 98109-1024, USA
^55^Department of Epidemiology, School of Public Health, University of Washington, Seattle, Washington 98195, USA
^56^International Hereditary Cancer Center, Department of Genetics and Pathology, Pomeranian Medical University, 70-115 Szczecin, Poland
^57^Faculty of Health and Medical Sciences, University of Copenhagen, 2200 Copenhagen, Denmark
^58^Department of Clinical Biochemistry, Herlev and Gentofte Hospital, Copenhagen University Hospital, Herlev, 2200 Copenhagen, Denmark
^59^Division of Clinical Epidemiology and Aging Research, German Cancer Research Center (DKFZ), D-69120, Heidelberg, Germany
^60^German Cancer Consortium (DKTK), German Cancer Research Center (DKFZ), D-69120 Heidelberg, Germany
^61^Division of Preventive Oncology, German Cancer Research Center (DKFZ) and National Center for Tumor Diseases (NCT), Im Neuenheimer Feld 460, 69120 Heidelberg, Germany
^62^Humangenetik Tuebingen, Paul-Ehrlich-Str 23, D-72076 Tuebingen, Germany
^63^The University of Texas M. D. Anderson Cancer Center, Department of Genitourinary Medical Oncology, 1515 Holcombe Blvd., Houston, TX 77030, USA
^64^Departments of Epidemiology & Population Health and of Medicine, Division of Oncology, Stanford Cancer Institute, Stanford University School of Medicine, Stanford, CA 94304 USA
^65^Department of Genetics, Portuguese Oncology Institute of Porto (IPO-Porto), 4200-072 Porto, Portugal
^66^Biomedical Sciences Institute (ICBAS), University of Porto, 4050-313 Porto, Portugal
^67^Cancer Genetics Group, IPO-Porto Research Center (CI-IPOP), Portuguese Oncology Institute of Porto (IPO-Porto), 4200-072 Porto, Portugal
^68^Department of Population Sciences, Beckman Research Institute of the City of Hope, 1500 East Duarte Road, Duarte, CA 91010, 626-256-HOPE (4673)
^69^Ghent University, Faculty of Medicine and Health Sciences, Basic Medical Sciences, Proeftuinstraat 86, B-9000 Gent
^70^Department of Surgery, Faculty of Medicine, University of Malaya, 50603 Kuala Lumpur, Malaysia
^71^Department of Urology, University of Washington, 1959 NE Pacific Street, Box 356510, Seattle, WA 98195, USA
^72^Institute of Human Genetics, University Medical Center Hamburg-Eppendorf, D-20246 Hamburg, Germany
^73^Molecular Medicine Center, Department of Medical Chemistry and Biochemistry, Medical University of Sofia, Sofia, 2 Zdrave Str., 1431 Sofia, Bulgaria
^74^Department of Oncology, Cross Cancer Institute, University of Alberta, 11560 University Avenue, Edmonton, Alberta, Canada T6G 1Z2
^75^Division of Radiation Oncology, Cross Cancer Institute, 11560 University Avenue, Edmonton, Alberta, Canada T6G 1Z2
^76^Molecular Endocrinology Laboratory, Department of Cellular and Molecular Medicine, KU Leuven, BE-3000, Belgium
^77^Division of Cancer Sciences, Manchester Cancer Research Centre, Faculty of Biology, Medicine and Health, Manchester Academic Health Science Centre, NIHR Manchester Biomedical Research Centre, Health Innovation Manchester, Univeristy of Manchester, M13 9WL
^78^The University of Surrey, Guildford, Surrey, GU2 7XH, UK
^79^Genomic Medicine Group, Galician Foundation of Genomic Medicine, Instituto de Investigacion Sanitaria de Santiago de Compostela (IDIS), Complejo Hospitalario Universitario de Santiago, Servicio Galego de Saúde, SERGAS, 15706, Santiago de Compostela, Spai
^80^University of California San Diego, Moores Cancer Center, Department of Family Medicine and Public Health, University of California San Diego, La Jolla, CA 92093-0012, USA
^81^Department of Urology, Erasmus University Medical Center, 3015 CE Rotterdam, The Netherlands
^82^"Exposome and Heredity", CESP (UMR 1018), Faculté de Médecine, Université Paris-Saclay, Inserm, Gustave Roussy, Villejuif
^83^Clinical Gerontology Unit, University of Cambridge, Cambridge, CB2 2QQ, UK
^84^Division of Epidemiology, Department of Internal Medicine, University of Utah School of Medicine, Salt Lake City, Utah 84132, USA
^85^George E. Wahlen Department of Veterans Affairs Medical Center, Salt Lake City, Utah 84148, USA
^86^Department of Laboratory Medicine and Pathology, Mayo Clinic, Rochester, MN 55905, USA
^87^Nuffield Department of Population Health, University of Oxford, United Kingdom
^88^Program in Genetic Epidemiology and Statistical Genetics, Department of Epidemiology, Harvard School of Public Health, Boston, MA, USA
^89^Division of Epidemiology, Department of Medicine, Vanderbilt University Medical Center, 2525 West End Avenue, Suite 800, Nashville, TN 37232 USA.
^90^International Epidemiology Institute, Rockville, MD 20850, USA
^91^Department of Epidemiology and Biostatistics, School of Public Health, Imperial College London, SW7 2AZ, UK
